# Wastewater Virus Detection Complements Clinical COVID-19 Testing to Limit Spread of Infection at Kenyon College

**DOI:** 10.1101/2021.01.09.21249505

**Authors:** Daniel Barich, Joan L. Slonczewski

## Abstract

In-person college instruction during the 2020 pandemic required effective and economical monitoring of COVID-19 prevalence. Kenyon College and the Village of Gambier conducted measurement of SARS-CoV-2 RNA from the village wastewater plant and from an on-campus sewer line. Wastewater RNA detection revealed virus prevalence leading to individual testing and case identification. Wastewater surveillance also showed when case rates had subsided, thus limiting the need for individual clinical testing. Overall, wastewater virus surveillance allows more targeted use of individual testing and increases community confidence in student population management.

## INTRODUCTION

The 2020 pandemic of COVID-19 led many colleges to close campus and replace in-person classes with remote instruction. In the fall, safe return to campus required widespread clinical testing of students for SARS-Cov-2 virus (1). For example, an analytical modeling study shows that clinical testing of all students every two days could successfully control the spread of infection (2). However, large-scale clinical testing incurs large costs, at a time when dedensified populations lead to loss of funding; and the overall uncertainties of disruption affect the morale of students and campus communities (3).

A measure proposed to supplement clinical testing is wastewater surveillance for SARS-CoV-2 viral RNA (4,5). Wastewater surveillance now provides a leading indicator of COVID-19 prevalence in hundreds of municipalities. In addition, universities report the use of wastewater virus monitoring to provide data equivalent to batch testing, as a way to detect new cases in college residences (6).

At Kenyon College, we included wastewater monitoring as an experimental measure in our plan for modified return to in-person instruction. We tested these predictions:

- Wastewater virus levels could alert us to the presence of viral infections before the appearance of symptoms and/or clinical test results. This information, together with other indicators, could lead to clinical testing and measures to curb virus spread.
- Decline of virus levels could indicate a decrease in virus prevalence, allowing a lower level of campus virus alert.
- Virus detection from a specific residential population could target our clinical testing for that population.
- Public reporting of wastewater surveillance could increase community confidence in the College management of student populations during the pandemic.

## METHODS AND SETTING

Kenyon College is a fully residential college located in central Ohio, in the Village of Gambier. In a typical year, Kenyon has 1,600 students in residence and the village has approximately 650 non-student residents. The college and village wastewater systems are served by a common wastewater treatment plant.

### Clinical testing

For the fall 2020 semester, the College invited 60 percent of students to be in residence in order to lower the density of campus and provide every student with a single room. When students arrived in August, all students and College employees underwent nasopharyngeal clinical testing by the CDC standard RT-PCR test for which the positive detection rate is 70%. Over the first two weeks of September, all students underwent two additional rounds of testing, followed by testing a quarter of all students weekly throughout the semester. By November 25, 7,600 clinical tests had been performed. All students then left campus, with the exception of 66 international students who remained in residence over the winter. Overall, in the fall semester clinical tests revealed 8 confirmed cases among 916 students in residence.

Clinical test results were obtained typically within 3-4 days of testing. All positive results from Kenyon College students and employees were reported to the Knox County Public Health department (KPH). KPH cooperated with our project by sharing dates for positive tests in the Village of Gambier. Thus we could compare wastewater virus with all data on Village cases, which includes residents both on and off Kenyon’s campus.

### Wastewater testing

In June, 2020, the College undertook a collaboration with the Village of Gambier to sample wastewater from the Gambier Wastewater Treatment Plant. At that time, there were estimated to be 700 residents in the village including 66 international Kenyon students. Over the last week of August, the student population increased to 920 (overall total, about 1600). The students remained till November 25, when the population again declined to 66. At that point all 66 remaining students were housed in a campus apartment complex whose wastewater enters a common line reachable by a manhole. Parallel sampling from the manhole commenced on November 30, on the same dates as the village wastewater plant.

Wastewater was sampled twice weekly, on Mondays and Thursdays at approximately 9:00 am. Grab samples were obtained from June 8 – 22, followed by 24-h composite samples thereafter. The NCA manhole samples included two grab samples (November 20 and December 3) followed by composite samples using a YSI PM-12 autosampler. All samples were shipped overnight to the laboratory, Source Molecular (now LuminUltra), Miami Lakes, Florida. At Source Molecular, viral RNA was analyzed by RT-PCR using the CDC primer set N1.

Approximately 30-200 ml sample was used for analysis. Sample was centrifuged as well as filtered using a negatively charged HA filter membrane. Both filtrate and centrifuged pellet were extracted for RNA. Matrix spike recovery was performed using heat-inactivated SARS-CoV-2 virus particles. From August 24 forward, the matrix spike was human coronavirus OC43.

RT-PCR results were obtained from the lab within 4-6 days after sample shipment. The data provided included Ct and RNA concentration for all samples and controls. Results reports were made available to the public by the Village of Gambier (https://villageofgambier.org/). Analysis of results was posted by Kenyon (http://biology.kenyon.edu/slonc/covid/Gambier_Wastewater_Report.html). The report site was public, and forty community members signed up for email updates.

## RESULTS AND DISCUSSION

We compared the results of wastewater virus testing with the clinical testing of individuals (Figure 1). Results in July and August suggested that in general when log_2_ RNA counts per liter were above 9 (512 copies/l) this level predicted a cluster of cases in the village. The College adopted a standard that whenever two virus results came in above 500 copies/l, a heightened virus alert would be called and all students tested. Such an event occurred on October 8 and 11, when we obtained two consecutive high virus levels. Subsequently all students were tested, and a “quiet period” was called to limit student activities. Clinical testing revealed one positive case (October 21) that would otherwise have been missed.

**Figure 1.**
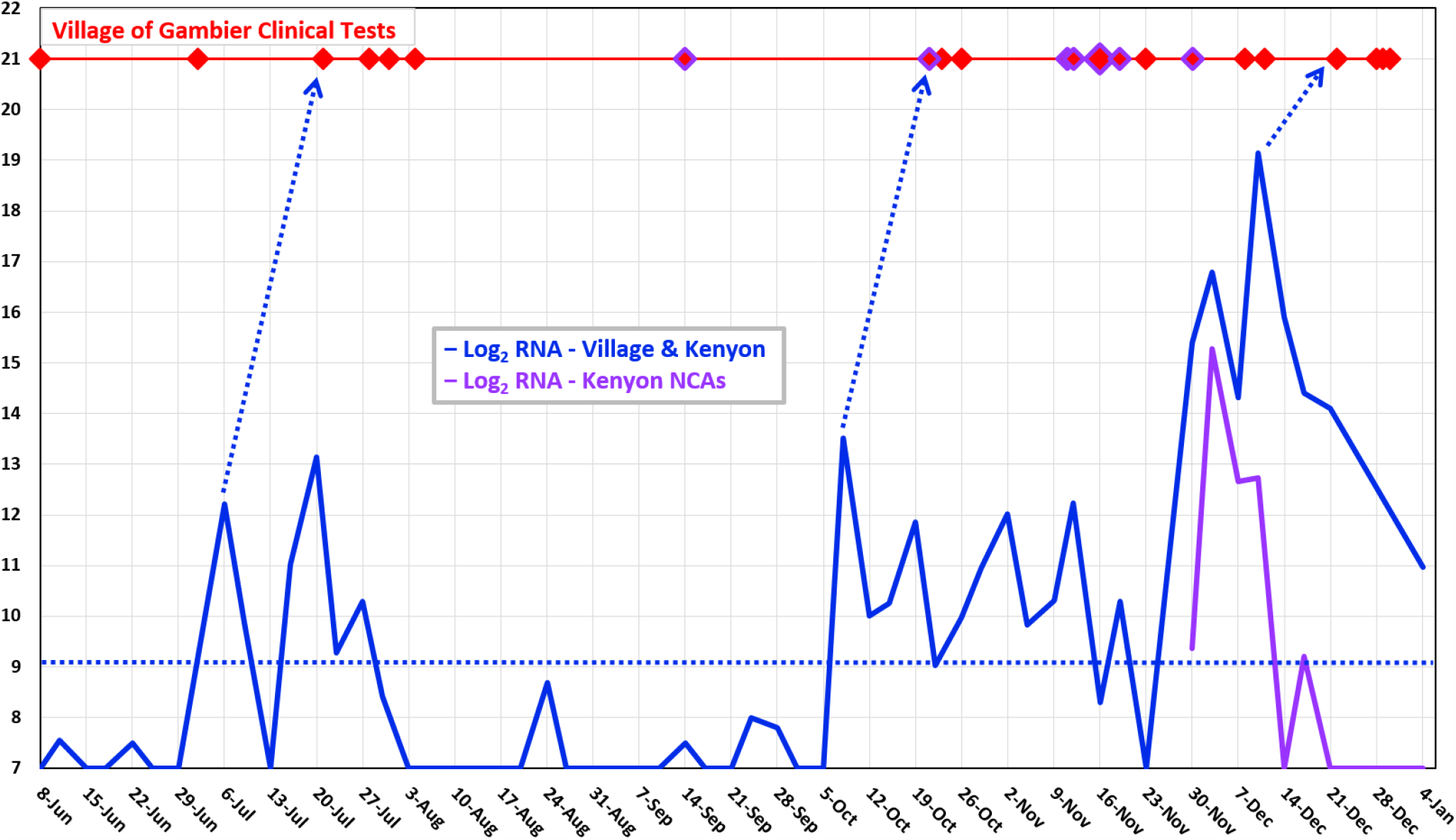
Wastewater SARS-CoV-2 RNA counts tracked with clinical testing. Blue line indicates log_2_ (viral RNA copies per liter) from the influent of the Village of Gambier wastewater treatment plant. Arrows indicate signal spike approximately two weeks ahead of a case cluster. Purple line indicates viral RNA from the North Campus Apartments manhole on Kenyon campus; counts are scaled down ten-fold. Red symbols indicate test dates for cases that were later reported positive to KPH. Purple outlined symbols indicate Kenyon students. Larger symbol represents two cases tested on one date. Horizontal dotted line indicates proposed level for case detection by wastewater RNA.

We estimated the lead time provided by the wastewater signal ahead of test results. A model was generated to test the relationship between twice-weekly wastewater RNA counts and daily clinical test reports from Knox Public Health. The model assumes 11 days of residual shedding post test date, with a two-day half-life as reported previously (7). A a dynamic range of 7 log_2_ units was assumed, based on the time of wastewater signal decay in July and October (Figure 1). The log_2_ values of lab-reported RNA counts per liter were interpolated to generate daily values. Positive clinical test dates were modeled using the date of testing followed by a series of virus shedding levels, starting with 2^6^ and declining by half every two days. Virus shedding levels per day were summed; days of zero shedding were indicated as one. The log_2_ RNA values were matched with the log_2_ estimates of individual virus shedding, using a Spearman’s rank correlation.

The correlation was performed with levels of offset (wastewater signal ahead of test dates) from 8-20 days. The peak value of correlation coefficient ρ was 0.53 at an offset of 14 days. Thus, the wastewater virus showed a tendency to peak about 14 days ahead of a cluster of confirmed COVID cases. In Figure 1, arrows indicate dates when clusters appeared to follow a wastewater signal peak.

When individual cases are detected clinically, they should track within a few days after the wastewater virus rises (7). The viral RNA in wastewater should rise steeply with the high shedding of virus that occurs early after infection (Figure 2). This early shedding is also the time of highest secondary transmission. A possible explanation for the two-week lead times we observed in July and in October is that primary cases of infection in the community were asymptomatic, and that only the secondary cases were detected and reported. Alternatively, cases tested clinically by RT-PCR were missed as false-negative results (30% expected).

**Figure 2.**
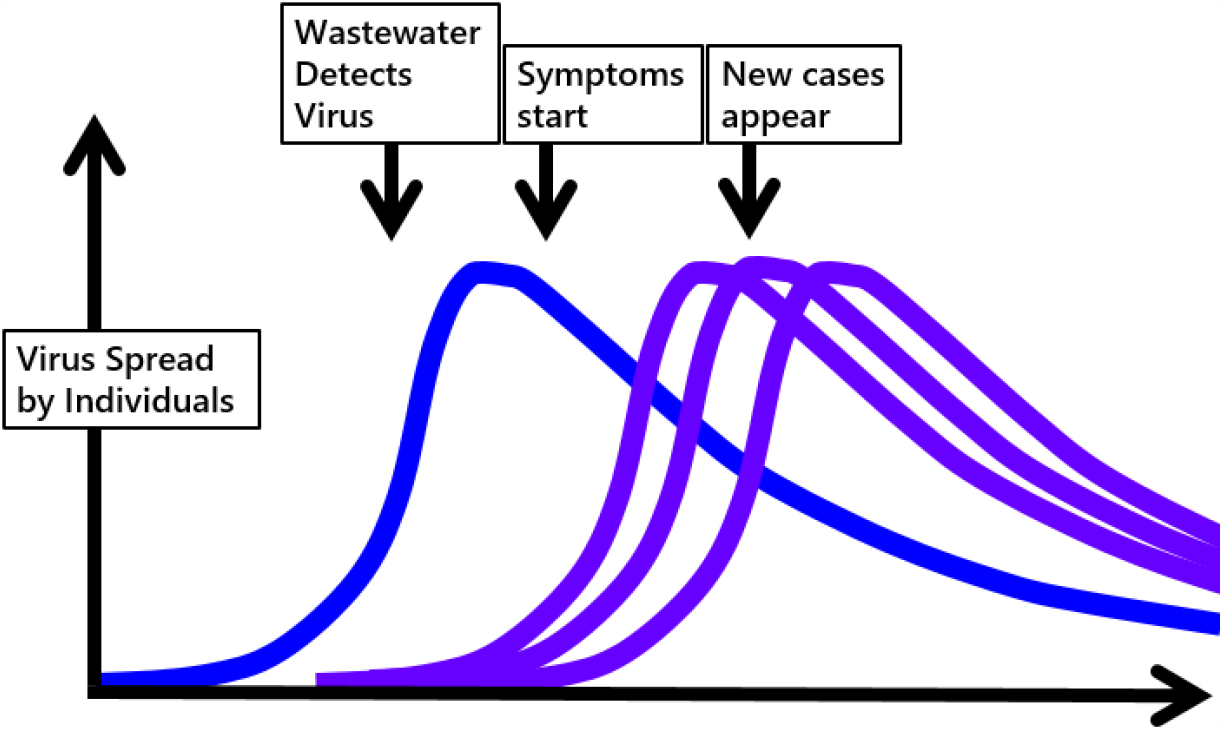
Model for relationship between wastewater SARS CoV-2 signal and COVID-19 infections. The wastewater RNA concentration starts to rise several days before symptoms occur. The first case may however be asymptomatic. Secondary infections occur early, becoming evident much later.

While the wastewater testing was useful for Kenyon’s COVID-19 surveillance, important limitations were noted. At least one positive case (September 14) was missed by the wastewater testing. Also, the testing from the village plant could not distinguish between cases on Kenyon’s campus versus non-Kenyon residents within the village. To focus testing on a student population, on November 30, Kenyon commenced testing from a manhole on campus. During that time, the manhole line received all wastewater from 66 students remaining on campus over the winter break. The manhole sampling revealed one possible case that may have been missed despite clinical testing of all 66 students (purple line in Figure 1). Thereafter, cases were reported in the village (population approximately 700, including the students) during a period when the campus manhole showed RNA levels below detection (indicating no student cases). Thus, the campus manhole testing appeared to successfully distinguish student cases from those off campus.

For all wastewater samples, the measure we used was the RNA copies per liter from RT-PCR provided directly by the testing lab. We considered whether these results required various kinds of normalization (Figure 3). In principle, the virus concentration must be normalized to wastewater volume, which varies with user population and rainwater infiltration. In the case of the Gambier plant, the source population size varied from 700 during summer and winter break, to a high of 1,600 during the fall semester. Nevertheless, the flow rate reported by the plant had a relatively modest effect on the virus signal (Figure 3, green line compared with blue line). During January, the signal from the Kenyon NCA manhole serves approximately one-tenth the population of the village plant (Figure 1). While no flow measurement was available, we represented the wastewater signal as one-tenth the size of the village signal.

**Figure 3.**
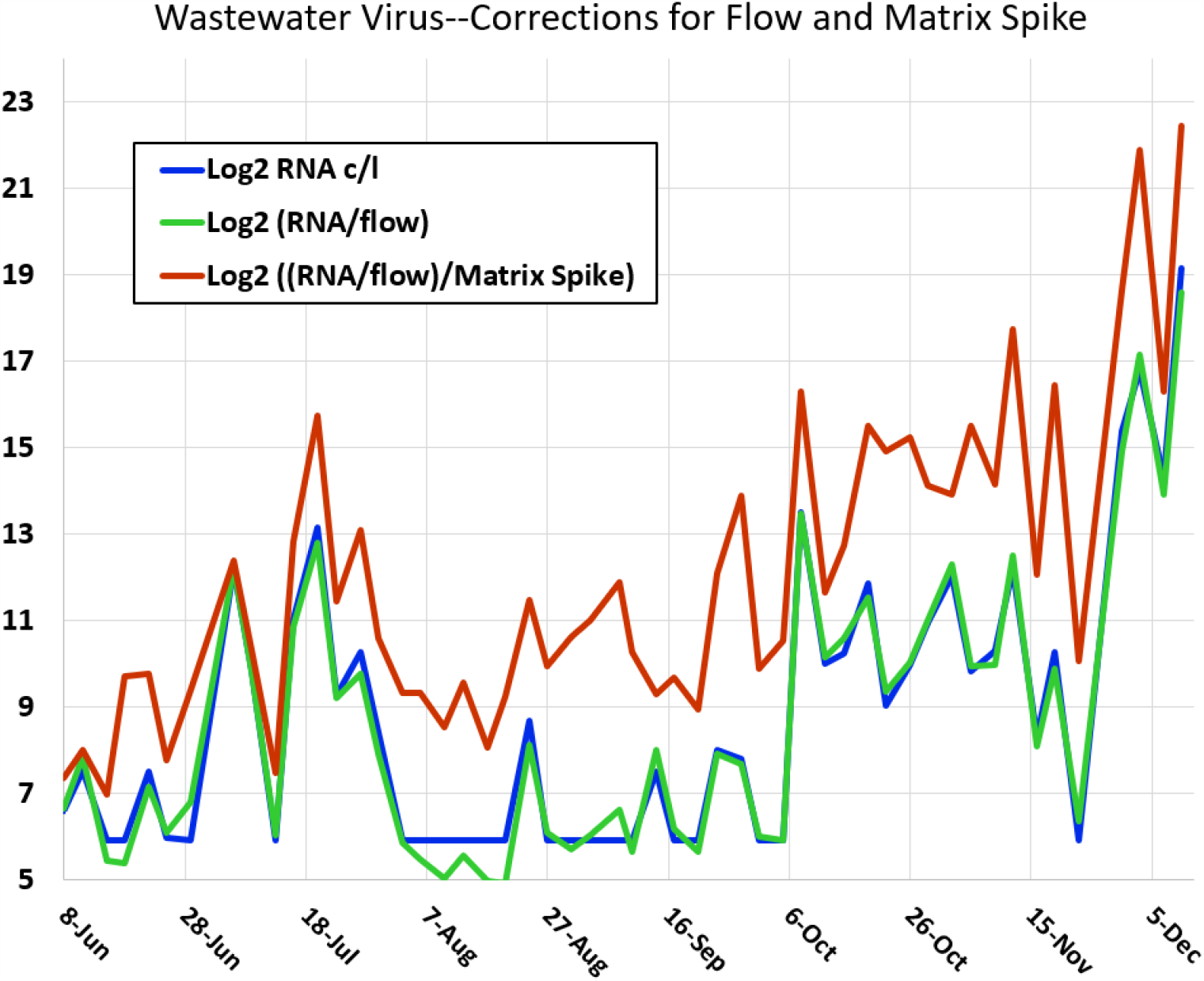
Wastewater signal correction factors. Blue line indicates log_2_ (viral RNA copies per liter) from the influent of the Village of Gambier wastewater treatment plant. Green line indicates viral concentration normalized to flow rate reported by the treatment plant. Brown line incidates flow-corrected viral concentration normalized to the percent detection of matrix spike.

We also attempted to normalize the reported RNA concentration to the detection efficiency for a spiked virus (Figure 3, brown line). The calculated result however did not improve the signal information and appeared to add noise. Spiked virus differs in character from the wastewater traces of viral RNA that are amplified by RT-PCR, so it may not provide an accurate measure of viral RNA signal efficiency.

Another means of normalization is to measure a host fecal marker such as crAssphage or pepper mild mottle virus (8). This fecal marker method was not available at the lab we used.

## CONCLUSIONS

In assessing our experiment, we emphasize that all wastewater results as well as clinical test results involve high levels of uncertainty. Nevertheless, given the community management tools available in 2020, our wastewater surveillance proved useful in these respects:

- Wastewater viral RNA levels enabled the College to increase clinical testing at times when the RNA signal was high, and to decrease priority of testing when the signal declined.
- Testing of a limited source (the NCA student apartment complex) enabled us to focus clinical testing on a limited population.
- Public reporting of wastewater surveillance drew considerable interest from the village and student community. There was increased confidence in the College and Village management of pandemic conditions, and increased support for measures such as mask wearing.

## Data Availability

All wastewater data are available from the Village of Gambier, and case testing dates are available from the Knox County Public Health department.

## ACKNOWLEDGMENTS

This study was supported by Kenyon College and by the Village of Gambier. We are grateful to the workers at the Gambier Wastewater Treatment Plant for their assistance. We thank the staff at Source Molecular (LuminUltra) for their prompt service and many valuable discussions. We thank members of the Ohio Department of Health and the Ohio universities working group on wastewater COVID-19 surveillance for their discussions. We especially thank Knox Public Health for their sharing of public case data.

